# Sarcopenic obesity and the risk of hospitalisation or death from COVID-19: findings from UK Biobank

**DOI:** 10.1101/2021.03.19.21253945

**Authors:** Thomas J. Wilkinson, Thomas Yates, Luke A. Baker, Francesco Zaccardi, Alice C. Smith

## Abstract

**Background:** Coronavirus disease□2019 (COVID□19) is an infectious disease caused by severe acute respiratory syndrome coronavirus 2 (SARS□CoV□2 virus). The role of skeletal muscle mass in modulating immune response is well documented. Whilst obesity is well-established as a key factor in COVID-19 infection and outcome, no study has examined the influence of both sarcopenia (low muscle mass) and obesity, termed ‘sarcopenic obesity’ on COVID-19 risk.

**Methods:** This study uses data from UK Biobank. Probable sarcopenia was defined as low handgrip strength. Sarcopenic obesity was mutually exclusively defined as the presence of obesity and low muscle mass (based on two established criteria: appendicular lean mass (ALM) adjusted for either: 1) height and 2) body mass index (BMI)). ‘Severe COVID-19’ was defined by a positive test result in a hospital setting or death with a primary cause reported as COVID-19. Fully adjusted logistic regression models were used to analyse the associations between sarcopenic status and severe COVID-19. This work was conducted under UK Biobank application number 52553.

**Results:** We analysed data from 490,301 UK Biobank participants. 2203 (0.4%) had severe COVID-19 infection. Individuals with probable sarcopenia were 64% more likely to have had severe COVID-19 infection (odds ratio (OR) 1.638; P<.001). Obesity increased the likelihood of severe COVID-19 infection by 76% (P<.001). Using either ALM index and ALM/BMI index to define low muscle mass, those with sarcopenic obesity were 2.6 times more likely to have severe COVID-19 (OR: 2.619; P<.001). Sarcopenia alone did not increase the risk of COVID-19.

**Conclusions:** Sarcopenic obesity may increase the risk of severe COVID-19 infection, over that of obesity alone. The mechanisms for this are complex but could be a result of a reduction in respiratory functioning, immune response, and ability to respond to metabolic stress.

## Introduction

Coronavirus disease□2019 (COVID□19), an infectious disease caused by severe acute respiratory syndrome coronavirus 2 (SARS□CoV□2 virus), reached pandemic status due to its infectivity and fatality [1]. As of the 15^th^ March 2021, in the UK there have been over 4 million confirmed cases resulting in over 120,000 deaths [2]. Age, sex, ethnicity, frailty, and the pre-existence of multiple comorbidities have been identified as important components associated with poor prognosis in COVID-19 [1, 3-5]. Obesity has been recognised as a risk factor in previous infectious outbreaks and is highly prevalent in individuals with COVID-19 [6]. Obesity is now well-established as a key factor of severe COVID-19 infection and mortality [7-12], likely due to adverse changes in pulmonary function and reductions in immune system function concomitant with excessive adiposity [13] which exacerbate the direct effects of COVID-19 induced pneumonia.

The impact of COVID-19 on sarcopenia, characterised by reduced muscle mass and muscle strength, has received substantial interest in the literature, although work has predominantly focused on how COVID-19 and its consequential social restrictions (e.g., physical activity) can cause acute loss of muscle and function [14, 15]. The role of skeletal muscle mass in modulating immune response and supporting metabolic stress responses are well documented. Patients with sarcopenia have been shown to have compromised intercostal muscle strength and respiratory function, which are detrimental in the treatment of severe pneumonia and acute respiratory distress syndrome [16], have higher incidence of community-acquired and in-hospital pneumonia, and are reported to have reduced ability to respond to systemic stress when facing acute infection, major surgeries, and other illness [14, 17, 18]. Currently, no study has investigated the relationship between sarcopenia and COVID-19 risk [14]. It is likely that individuals with sarcopenia respond poorly to infection with COVID-19 because of impaired immune potential.

The presence of both sarcopenia and obesity is termed ‘sarcopenic obesity’ [19] and has been associated with increased risk of disability, institutionalization, mortality, and metabolic diseases compared to sarcopenia or obesity alone [20, 21]. Consequently, given the potential adverse consequences of both obesity and sarcopenia on COVID-19 risk, it may be reasonable to expect that sarcopenic obesity could result in more serious prognosis and infection risk. To our knowledge, there have been no community-based studies on the association between sarcopenic status and risk of COVID-19 infection. In this study we aimed to compare the association between sarcopenic status, alone and in combination with obesity, and severe COVID-19 infection resulting in hospital admission or death in UK Biobank.

## Methods

### Data source

This study uses data from UK Biobank. Over 500,000 participants, aged 37 to 73□years from the general population, were recruited into UK Biobank study between March 2006 and December 2010. Participants attended one of 22 assessment centres across the UK where they completed a touch-screen questionnaire, had physical measurements taken, and provided biological samples, as described in detail elsewhere [22]. UK Biobank was approved by the North West Multi-Centre Research Ethics Committee (11/NW/0382). All participants provided written informed consent to participate in the UK Biobank study. This work was conducted under the UK Biobank application number 52553.

### Exposure

Our exposure of interest was sarcopenic obesity status as reported at baseline. We undertook our analysis using two different sarcopenia criteria proposed by the European Working Group of Sarcopenia in Older People (EWGSOP2) and Foundation for the National Institutes of Health (FNIH) Sarcopenia Project. The role of dynapenia, a condition characterized by an age-related decline in muscle strength, is now considered a principal determinant of ‘probable sarcopenia’ [23]. In our sample, probable sarcopenia was defined as low handgrip strength (HGS) (<16kg in females, <27kg in males). The maximum HGS score was taken from the highest value attained in either the right or left hand.

Low muscle mass was defined as an appendicular lean mass (ALM) below specific cut-off indices. ALM was derived from the sum of fat-free mass (FFM) of the arms and legs taken from bioelectrical impedance analysis (BIA) data assessed during baseline assessment visit. ALM was calculated using a previous published equation [24], which estimated ALM from the appendicular FFM values: ALM (kg) = (0.958 × [Appendicular FFM (kg)]) − (0.166 × G) − 0.308, with G taking value 0 if female and 1 if male.

Sarcopenia was defined as either:

1. ALM index (ALM/height^2^) <7.26 kg/m^2^ for males and <5.45 kg/m^2^ for females as per EWGSOP2 criteria [23].
2. ALM/BMI index (ALM/BMI) <0.789 in males and <0.512 in females as per FNIH Sarcopenia Project criteria [25].

Sarcopenic obesity was mutually exclusively defined as the presence of obesity and low muscle mass (using both ALM index definitions). Obesity was defined as excessive body fat % (BF%) using cut-offs of >25% in males and >35% in females [21]. We defined obesity using adiposity (i.e., BF%) rather than BMI which can be confounded by the inclusion of lean mass. A ‘normal’ reference group consisted of those without sarcopenia, obesity, or sarcopenic obesity as per the definitions above.

### Outcomes

Public Health England (PHE) provided the SARS-CoV-2 test results, including the specimen date, origin (evidence that the individual was an inpatient or not), and result (positive or negative) of the test. Latest test result data were available up until 2^nd^ February 2021. Records were linked to inpatient Hospital Episode Statistics and national mortality registers (latest mortality data were available up to 16^th^ February 2021). As per previous studies [4, 26], the outcome of interest was ‘severe COVID-19’. This was defined by a positive test result for SARS-CoV-2 in a hospital setting (i.e., participants whose tests were taken while an inpatient) or death with a primary cause reported as COVID-19 (ICD-10 code U07.1) in accordance with WHO guidance [27]. Participants were excluded from the dataset if they were not alive during the pandemic (i.e., died before 11^th^ March 2020, the date pandemic declared by WHO).

### Confounders

Covariates of interest included: current age, which was calculated on 16^th^ March 2020, the first available day for linkage used in this analysis; sex; ethnicity (defined as White or non-White); area-based socioeconomic status (deprivation) derived from the postcode of residence, using the Townsend deprivation index (a composite measure of deprivation based on unemployment, non-car ownership, non-home ownership, and household overcrowding; a negative value represents high socioeconomic status); and total number of cancer and non-cancer reported illnesses.

### Statistical analysis

Analysis was based on a whole population level approach as described previously for UK Biobank analysis [28], with ‘severe COVID-19’ cases compared to the remaining UK Biobank population. Fully adjusted logistic regression models were used to analyse the associations between sarcopenic status (defined using either criteria) and severe COVID-19. A sensitivity analysis examining the association of each ALM index individually was also performed. The results are reported as odds ratios (ORs) with their 95% confidence intervals (95%CI).

## Results

### Participant characteristics

In total, data (with exposure and confounders) were available for 490,301 participants, of which 2203 (0.4%) had severe COVID-19 infection. Characteristics of participants, stratified by COVID-19 status, are reported in **Table 1**. The median age was 70 years, with approximately 50% of the cohort male. The majority of participants were White.

**Table 1.** Basic participant characteristics stratified by severe COVID-19 status.

|  | **No COVID-19 associated admission or death** | **Severe COVID-19 infection** |
| --- | --- | --- |
| Total n | 488,098 | 2203 |
| Age (years) | 70.0 (13.0) | 70.0 (16.0) |
| Sex (male n, %) | 225,559 (46%) | 1146 (52%) |
| *Ethnicity* |  |  |
| White | 462,460 (95%) | 1986 (90%) |
| Non-White | 25,638 (5%) | 217 (10%) |
| Number of cancer illnesses | 1.0 (3.0) | 2.0 (2.0) |
| Number of non-cancer illnesses | 0.0 (0.0) | 0.0 (0.0) |
| Townsend deprivation index | -2.2 (4.2) | -1.2 (5.0) |
| Probable sarcopenia | 26,444 (5%) | 189 (9%) |
| *Low muscle mass* |  |  |
| Low ALM/height<sup>2</sup> index | 8296 (2%) | 25 (1%) |
| Low ALM/BMI index | 8223 (2%) | 70 (3%) |
| Either index | 9267 (2%) | 75 (4%) |
| Obesity | 270,063 (57%) | 1501 (70%) |

*Data shown as median and inter-quartile range (IQR) or n (%)*

### Probable sarcopenia and risk of severe COVID-19 infection

Probable sarcopenia was present in 189 (8.6%) of those with severe COVID-19, compared to 26,444 (5.4%) in those without COVID-19-associated admission or death. Individuals with probable sarcopenia were 64% more likely to have had severe COVID-19 infection (adjusted OR 1.638 (95% CI: 1.411 to 1.903); P<.001), compared to those without probable sarcopenia (**Figure 1** and **Supplementary Material Table S1 and S2**). There was no significant effect of obesity on the association between probable sarcopenia and risk of severe COVID-19 (P=.839) (**Table S3**).

**Figure 1.**
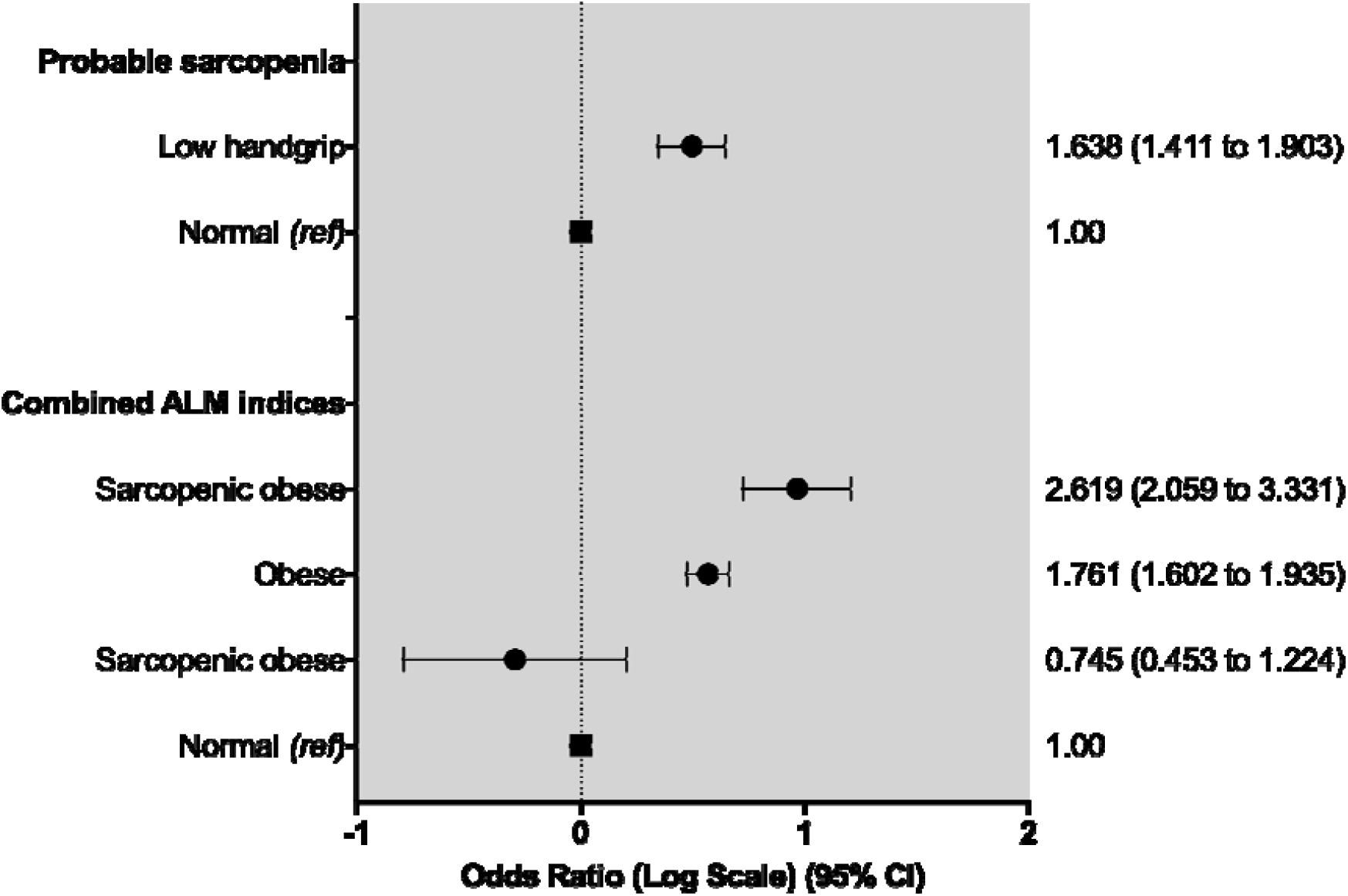
Odds ratios for risk of severe COVID-19 infection across sarcopenia status. Data presented as odds ratio and 95% confidence intervals (95% CI). Adjusted for current age, sex, ethnicity, Townsend deprivation index, number of cancer and non-cancer illnesses. Probable sarcopenia was defined as low handgrip strength (<16kg in females, <27kg in males); sarcopenic obese defined a presence of obesity and sarcopenia (defined as either low muscle using ALM/height index or ALM/BMI index); a ‘normal’ reference group consisted of those without sarcopenia, obesity, or sarcopenic obesity

### Sarcopenic obesity and risk of severe COVID-19 infection

Using either index to define low muscle mass, data were available for 478,683 participants, of which 2133 (0.4%) had severe COVID-19 infection. In participants with severe COVID-19 infection, 616 (28.9%) had no sarcopenia, obesity, or sarcopenic obesity (compared to 199,646 (41.9%) in those without severe COVID-19. Sixteen (0.8%) severe COVID-19 cases had sarcopenia (versus 6964, 1.5%), 1426 (66.9%) were obese (versus 260,673, 54.7%), and 75 (3.5%) had sarcopenic obesity (versus 9267, 1.9%; **Table S4**).

Obese individuals were 1.7 times more likely to have severe COVID-19 infection (adjusted OR 1.761 (95% CI: 1.602 to 1.935); P<.001). Individuals with sarcopenic obesity were 2.6 times more likely to have severe COVID-19 (adjusted OR: 2.880 (95% CI: 2.248 to 3.691); P<.001) (**Figure 1** and **Table S5**). Sarcopenia alone did not increase the risk of severe COVID-19.

### Sarcopenic obesity and risk of severe COVID-19 infection using ALM index and ALM/BMI index

A sensitivity analysis of both ALM index and ALM/BMI index individually did not change the association with severe COVID-19 (data shown in **S6-S9, Figure S1**)

## Discussion

Although evidence suggests that obesity is a key risk factor for COVID-19 risk, no data exists on the association of sarcopenia and COVID-19, and also, importantly, when obesity occurs in the presence of sarcopenia (or low muscle mass), termed ‘sarcopenic obesity’. Using UK Biobank, we examined the association with different measures of sarcopenia and sarcopenic obesity with severe COVID-19 infection. We found: 1) individuals with probable sarcopenia (defined as low HGS) were at higher risk of severe COVID-19 compared to those without probable sarcopenia; 2) sarcopenic obesity significantly increased the likelihood of severe COVID-19 above that of obesity alone (regardless of low muscle mass definition).

It is well-established that obesity is a significant risk factor in severe COVID-19 infection and COVID-19 related mortality [7-12]. The link between obesity and COVID-19 infection is multifaceted: obesity is associated with decreased expiratory reserve volume, functional capacity, and respiratory system compliance. In patients with increased abdominal obesity, pulmonary function is further compromised in supine patients by decreased diaphragmatic excursion, making ventilation more difficult. Furthermore, increased inflammatory cytokines associated with obesity may contribute to the increased morbidity associated with obesity in COVID□19 infections [13]. Our data supports the previous work showing obesity remains a significant factor in COVID-19 infection.

Sarcopenia is characterized by reduced muscle mass and muscle strength. Although the exact criteria to define it differ, reduced skeletal muscle mass is a fundamental feature across all definitions. There is robust evidence for an impaired immune response in patients with sarcopenia, including a higher incidence of community-acquired and in-hospital pneumonia, and an increased risk of infectious complications following surgeries [14, 17, 18]. A key mechanism underlying the impaired immunity in individuals with sarcopenia refers to the abnormal myokines, such as interleukin (IL)-15, IL-17, and IL-6, which modulate the proliferation and function of both innate and adaptive immune cells [29]. During severe infection, skeletal muscle is catabolized to provide the immune system, liver, and gut with amino acids, especially glutamine [30]; patients with sarcopenia have a decreased availability of such protein mobilization. Furthermore, compromised intercostal muscle strength and respiratory function could further exacerbate a failure in ventilatory function. In the context of COVID-19 risk, therefore, it is likely skeletal muscle has an important role in respiratory functioning, modulating immune response, and supporting metabolic stress [14].

Given the low prevalence of mutually exclusive sarcopenia (i.e., individuals with just low muscle mass) in UK Biobank [24], it was difficult to ascertain the independent effect of sarcopenia alone on severe COVOD-19 risk. However, we found that there was a greater number of individuals with sarcopenic obesity, which was present in over 9000 individuals when sarcopenia was defined using either index. In particular, the prevalence of sarcopenic obesity was greater when using ALM adjusted for BMI (recommended as a more valid index of muscle mass above ALM adjusted for only height [25, 31]). We found sarcopenic obesity increased the odds of severe COVID-19 infection approximately 3 times, compared to those who were just obese (1.8 times). Given the detrimental independent effects of low muscle mass and obesity, this result in perhaps unsurprising, although highlights the severe detrimental effects of sarcopenic obesity. It is important to note that the risk of COVID-19 did not significantly change across either ALM index to define low muscle mass.

The interaction between sarcopenia, obesity, and sarcopenic obesity with COVID-19 is likely bidirectional. COVID-19 could be a risk factor for the incidence and progression of sarcopenic obesity because of the reduced physical activity and inadequate diet (e.g., low protein intake) caused by social isolation and restrictions in mobility. The inflammatory reaction caused by COVID-19 may also exacerbate metabolic stress and muscle catabolism [14], although we cannot infer this in our cohort here. Therapeutic approaches targeting skeletal muscle and adiposity should form a fundamental part in the treatment of COVID-19. Physical activity, especially resistance exercise, can help increase muscle mass and reduce body fat, whilst appropriate nutritional management (i.e., a balanced nutritional formula with high-quality protein – rich in leucine) can help promote muscle synthesis [14].

Whilst UK Biobank offered a large cohort of individuals with linked data to both COVID-19 testing and outcomes, it is limited by low initial response rate (5.5%) and evidence of a ‘healthy responder’ bias [32]. This ‘healthy responder’ bias may explain the low prevalence of sarcopenia as previously described [24]. Sarcopenic obesity status was also taken from baseline assessments which were performed a decade ago. Given that sarcopenia in particular is likely to worsen with age, the results presented here could underestimate the current sarcopenia prevalence, and therefore overestimate the effect on COVID-19 risk.

In conclusion, the presence of sarcopenic obesity may increase the risk of severe COVID-19 infection, over that of obesity alone. The mechanisms for this association are complex but could be a result of a reduction in respiratory functioning, immune response, and ability to respond to metabolic stress. Therapeutic approaches targeting both skeletal muscle and adiposity should form a key part in the treatment of COVID-19.

## Data Availability

Data included in this article available through application to UK Biobank.

## Acknowledgments

The authors of this manuscript certify that they comply with the ethical guidelines for authorship and publishing in the Journal of Cachexia, Sarcopenia and Muscle [33].

