## Supplementary material for "Sarcopenic obesity and the risk of hospitalisation or death from COVID-19: findings from UK Biobank"

**Probable sarcopenic and risk of severe COVID-19 infection**

**Table S1. Probable sarcopenia stratified by severity of COVID-19 infection**

|  | **No COVID-19 associated admission or death (n=488,098)** | **Severe COVID-19 infection (n=2203)** |
| --- | --- | --- |
| Normal (n=463,668) | 461,654 (94.6%) | 2014 (91.4%) |
| Probable sarcopenia (n=26,633) | 26,444 (5.4%) | 189 (8.6%) |

**Table S2. Association of probable sarcopenia with risk of severe COVID-19 infection**

|  | **OR** | **95%CI Upper** | **95%CI Lower** | **P** |
| --- | --- | --- | --- | --- |
| Normal (Ref.) | 1.000 | - | - | - |
| Probable sarcopenia | 1.638 | 1.411 | 1.903 | <.001* |

Adjusted for age, sex, ethnicity, Townsend deprivation index, number of cancer and non-cancer illnesses

**Table S3. Association of probable sarcopenia with risk of severe COVID-19 infection stratified by obesity status**

|  | **OR** | **95%CI Upper** | **95%CI Lower** | **P** |
| --- | --- | --- | --- | --- |
| **Obese** |  |  |  |  |
| Normal (Ref.) | 1.000 | - | - | - |
| Probable sarcopenia | 1.449 | 1.213 | 1.730 | <.001* |
| **Non-obese** |  |  |  |  |
| Normal (Ref.) | 1.000 | - | - | - |
| Probable sarcopenia | 1.573 | 1.132 | 2.185 | <.001* |

Adjusted for current age, sex, ethnicity, Townsend deprivation index, number of cancer and non-cancer illnesses

**Sarcopenic obesity and risk of severe COVID-19 infection using either ALM indices**

**Table S4. Sarcopenic status stratified by severe COVID-19 infection using either ALM indices**

|  | **No COVID-19 associated admission or death (n=476,550)** | **Severe COVID-19 infection (n=2133)** |
| --- | --- | --- |
| Normal (n=200,262) | 199,646 (41.9%) | 616 (28.9%) |
| Sarcopenic (n=6980) | 6964 (1.5%) | 16 (0.8%) |
| Obese (BF%) (n=265,408) | 260,673 (54.7%) | 1426 (66.9%) |
| Sarcopenic obese (n=9358) | 9267 (1.9%) | 75 (3.5%) |

Sarcopenic obese defined a presence of obesity and sarcopenia (defined as either low muscle using ALM/height index or ALM/BMI index)

**Table S5. Association of sarcopenic status with risk of severe COVID-19 infection using either ALM indices**

|  | **OR** | **95%CI Upper** | **95%CI Lower** | **P** |
| --- | --- | --- | --- | --- |
| Normal (Ref.) | 1.000 | - | - | - |
| Sarcopenic | 0.745 | 0.453 | 1.224 | .245 |
| Obese (body fat %) | 1.761 | 1.602 | 1.935 | <.001 |
| Sarcopenic obese | 2.619 | 2.059 | 3.331 | <.001 |

Adjusted for current age, sex, ethnicity, Townsend deprivation index, number of cancer and non-cancer illnesses.

**Sarcopenic obesity and risk of severe COVID-19 infection using ALM index**

**Table S6. Sarcopenic status stratified by severe COVID-19 infection using ALM index**

|  | **No COVID-19 associated admission or death (n=476,551)** | **Severe COVID-19 infection (n=2133)** |
| --- | --- | --- |
| Normal (n=200,499) | 199,883 (41.9%) | 616 (28.8%) |
| Sarcopenic (ALM index) (n=6743) | 6727 (1.4%) | 16 (0.8%) |
| Obese (BF%) (n=269,928) | 268,436 (56.3%) | 1492 (69.9%) |
| Sarcopenic obese (n=1514) | 1505 (0.3%) | 9 (0.4%) |

Sarcopenic obese defined a presence of obesity and sarcopenia (defined as low ALM index)

Using ALM index to define low muscle mass, data were available for 478,684 participants, of which 2133 (0.4%) had severe COVID-19 infection. In participants with severe COVID-19 infection, 616 (28.8%) had no sarcopenia, obesity, or sarcopenic obesity (compared to 199,883 (42%) in those without severe COVID-19. Sixteen (0.8%) of severe COVID-19 cases had sarcopenia (versus 6727, 1.4% in those without severe COVID-19), 1492 (69.9%) were obese (versus 268,436, 56.3%), and 9 (0.4%) had sarcopenic obesity (versus 1492, 0.3%).

**Table S7. Association of sarcopenic status with risk of severe COVID-19 infection using ALM index**

|  | **OR** | **95%CI Upper** | **95%CI Lower** | **P** |
| --- | --- | --- | --- | --- |
| Normal (Ref.) | 1.000 | - | - | - |
| Sarcopenic (ALM index) | 0.772 | 0.470 | 1.269 | .307 |
| Obese (body fat %) | 1.804 | 1.642 | 1.981 | <.001* |
| Sarcopenic obese | 1.940 | 1.003 | 3.754 | .049* |

Adjusted for current age, sex, ethnicity, Townsend deprivation index, number of cancer and non-cancer illnesses. Sarcopenic only individuals (ALM index) were removed from the analysis due to the small number of cases.

Compared to those without obesity or sarcopenia, obesity alone increased the likelihood of severe COVID-19 infection by 80% (adjusted OR: 1.804 (95% CI: 1.642 to 1.981); P<.001). The presence of both sarcopenia and obesity increased severe COVID-19 risk further; those with sarcopenic obesity were 1.9 times more likely to have severe COVID-19 (adjusted OR: 1.940 (95% CI: 1.003 to 3.754); P=.049). Sarcopenia, on its own, was not associated with an increased risk of severe COVID-19).

**Sarcopenic obesity and risk of severe COVID-19 infection using ALM/BMI index**

**Table S8. Sarcopenic status stratified by severe COVID-19 infection using ALM/BMI index**

|  | **No COVID-19 associated admission or death (n=476,550)** | **Severe COVID-19 infection (n=2133)** |
| --- | --- | --- |
| Normal (n=206,968) | 206,336 (43.3%) | 632 (29.6%) |
| Sarcopenic (ALM/BMI index) (n=274) | 274 (0.1%) | 0 (0%) |
| Obese (BF%) (n=263,437) | 262,006 (54.9%) | 1431 (67.1%) |
| Sarcopenic obese (n=8001) | 7934 (1.7%) | 70 (3.3%) |

Sarcopenic obese defined a presence of obesity and sarcopenia (defined as low ALM/BMI index)

Using ALM/BMI index to define low muscle mass, data were available for 478,683 participants, of which 2133 (0.4%) had severe COVID-19 infection. In participants with severe COVID-19 infection, 632 (29.6%) had no sarcopenia, obesity, or sarcopenic obesity (compared to 206,336 (43%) in those without severe COVID-19. No (0%) severe COVID-19 cases had sarcopenia (versus 278, 0.1%), 1431 (67.1%) were obese (versus 262,006, 54.9%), and 70 (3.3%) had sarcopenic obesity (versus 7934, 1.7%).

**Table S9. Association of sarcopenic status with risk of severe COVID-19 infection using ALM/BMI index**

|  | **OR** | **95%CI Upper** | **95%CI Lower** | **P** |
| --- | --- | --- | --- | --- |
| Normal (Ref.) | 1.000 | - | - | - |
| Obese (body fat %) | 1.783 | 1.624 | 1.958 | <.001* |
| Sarcopenic obese | 2.880 | 2.248 | 3.691 | <.001* |

Adjusted for current age, sex, ethnicity, Townsend deprivation index, number of cancer and non-cancer illnesses

Sarcopenic only individuals (ALM/BMI index) were removed from the analysis due to the small number of cases.

Obesity increased the likelihood of severe COVID-19 infection by 79% (adjusted OR 1.783 (95% CI: 1.624 to 1.958); P<.001). Sarcopenic obesity further increased this risk, with individuals 2.9 times more likely to have severe COVID-19 (adjusted OR: 2.880 (95% CI: 2.248 to 3.691); P<.001) (Figure 1 and Table S7). Sarcopenic only individuals (ALM/BMI index) were removed from the analysis due to the small number of cases.

**Figure S1. Sarcopenic status stratified by severe COVID-19 infection**

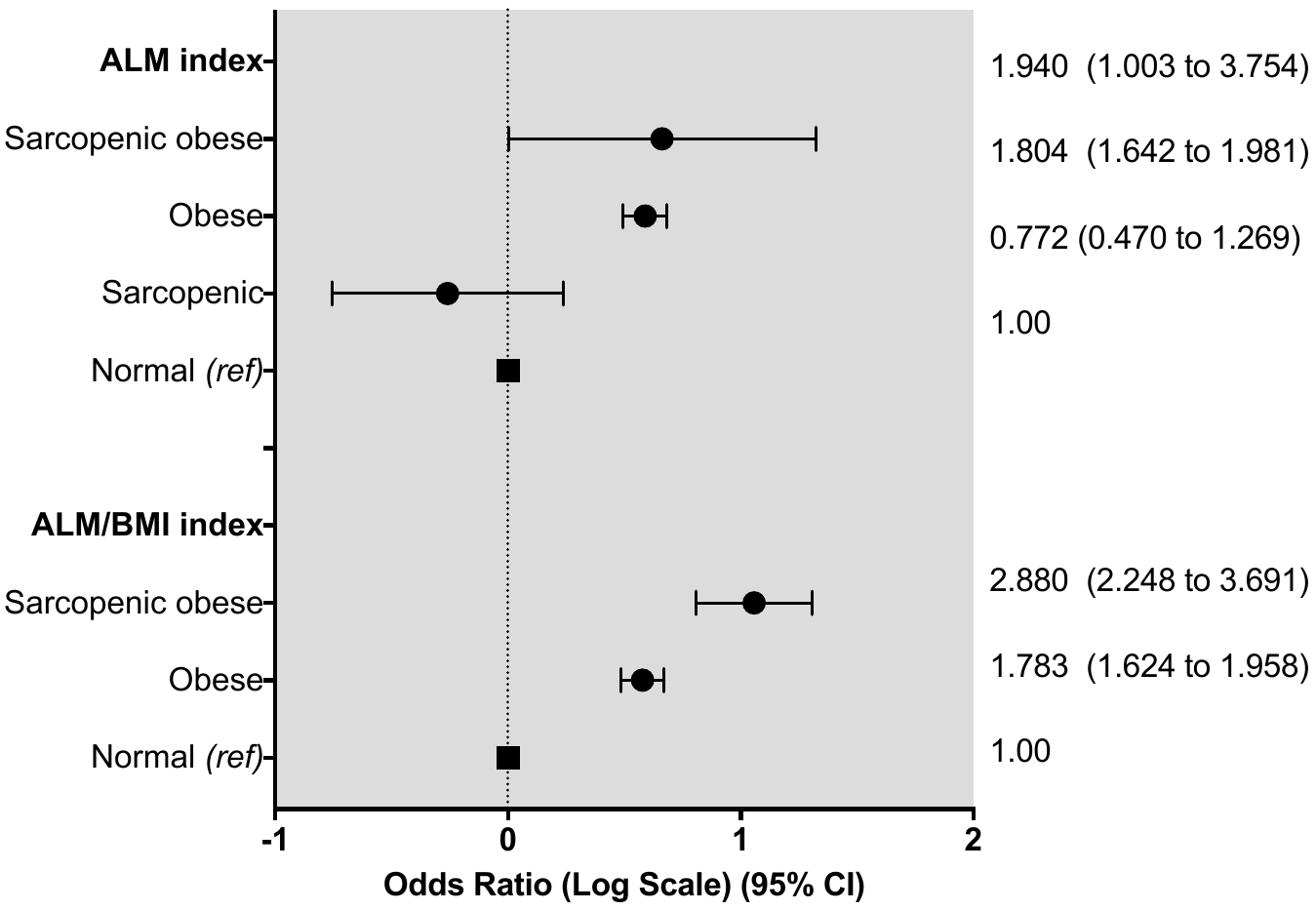
